# The Impact of Viral and Host Factors on the Influenza A Virus Transmission Bottleneck

**DOI:** 10.64898/2025.12.02.25341469

**Authors:** Kathryn C. Krupinsky, Emily E. Bendall, Yuwei Zhu, Melissa S. Stockwell, Huong Q. Nguyen, Jennifer K. Meece, Yvonne Maldonado, Katherine D. Ellingson, Karen Lutrick, Edwin J. Asturias, Suchitra Rao, Natalie M. Bowman, Melissa Rolfes, Jessica E. Biddle, Alexandra Mellis, Jonathan E. Schmitz, James D. Chappell, Natasha B. Halasa, William J. Fitzsimmons, Emily T. Martin, Carlos G. Grijalva, H. Keipp Talbot, Adam S. Lauring

**Affiliations:** Department of Microbiology & Immunology, University of Michigan, Ann Arbor, MI, USA; Department of Biostatistics, Vanderbilt University Medical Center, Nashville, TN, USA; Department of Pediatrics and Department of Population and Family Health, Columbia University Irving Medical Center, New York, NY, USA; Marshfield Clinic Research Institute, Marshfield, WI, USA; Department of Pediatrics, Stanford University, Palo Alto, CA, USA; Department of Epidemiology & Biostatistics, University of Arizona, Tucson, AZ, USA; Department of Family and Community Medicine, University of Arizona, Tucson, AZ, USA; Department of Pediatrics, University of Colorado Anschutz Medical Campus, Aurora, CO USA; Division of Infectious Diseases, University of North Carolina, Chapel Hill, NC USA; Influenza Division, Centers for Disease Control and Prevention, Atlanta, GA USA; Department of Pathology, Vanderbilt University Medical Center, Nashville, TN, USA; Department of Pediatrics, Vanderbilt University Medical Center, Nashville, TN; Division of Infectious Diseases, Department of Internal Medicine, University of Michigan, Ann Arbor, MI, USA; Department of Epidemiology, University of Michigan, Ann Arbor, MI, USA; Department of Health Policy, Vanderbilt University Medical Center, Nashville, TN, USA; Department of Medicine, Vanderbilt University Medical Center, Nashville, TN, USA

## Abstract

**Background:** Transmission bottlenecks are defined by the number of unique virions or genotypes that establish an infection, and they restrict the viral diversity that passes from one infected host to another. They have a major impact on the spread of mutations in a population. Previous work has identified a tight transmission bottleneck for seasonal influenza A virus (IAV) based on an analysis of 43 household pairs, largely from a single A(H3N2) predominant season. While many viral and host factors are known to influence IAV transmission in households, their impact on the transmission bottleneck is not clear.

**Methods:** Nasal swabs were collected daily from IAV infected individuals enrolled in two case-ascertained U.S. household transmission studies, FluTES (2017/2018 through 2019/2020 seasons) and RVTN (2021/2022 season). Viruses were sequenced in duplicate, and intrahost single nucleotide variants (iSNV) were identified at a 0.5% frequency threshold using a benchmarked pipeline with >99.99% specificity for mutations present in both replicates. Transmission pairs were defined based on co-residence, test date, and genetic distance. For each possible transmission pair, the bottleneck was estimated using a beta binomial and a clonal mutation model.

**Results:** We sequenced 567 samples from 319 individuals and 102 households in duplicate. Based on epidemiologic linkage and a sequence-based cut-off, we defined 56 transmission pairs for the beta binomial model and 60 transmission pairs for the clonal mutation model. Across all pairs, we identified a transmission bottleneck of 1 both using the beta-binomial model (CI 1, 1) and the clonal mutation model (exact estimate: 0.91, CI: 0.91, 0.97). In our cohort, influenza season, subtype, and host factors (influenza vaccination status, sex, and age) did not alter the transmission bottleneck.

**Conclusions:** IAV is subject to a tight genetic bottleneck during transmission, which limits the onward propagation of newly arising nucleotide variants. Tight bottlenecks appear to be intrinsic to the transmission process, as host and viral factors within households do not affect its size.

## INTRODUCTION

Many respiratory viruses, and influenza virus in particular, undergo antigenic drift – the selection of newly arising strains that escape host immunity. This complicates long-term within-host immune memory as well as disease management and control (1,2). Rapid evolution can also potentially lead to new strains that differ in their transmissibility and pathogenicity (3–5). While viral nucleotide variants are typically identified through population-level surveillance, each nucleotide variant ultimately arises from mutations that occur during a host’s infection. Whether or not a given nucleotide variant reaches a secondary host and subsequently propagates within the population is a highly complex process. One important factor is the size of the transmission bottleneck, or the number of unique variants that establish a new infection (6). When a transmission bottleneck is tight, few nucleotide variants transfer from one host to another, and the virus is largely identical across a transmission chain. Conversely, when a transmission bottleneck is loose, a larger number of nucleotide variants transfer from one host to another, and a combination of rare and common nucleotide variants establish infection in the next host. Because bottlenecks determine which mutations propagate along a transmission chain, they are important determinants of evolutionary rate (7,8).

Given technical limitations in capturing the precise moment of infection establishment, it is difficult to observe transmission bottlenecks. In experimental systems, infections of animals with barcoded viruses and subsequent barcode quantification have been used to directly determine bottleneck size (9). However, in observational human studies, we rely on inference methods to determine bottleneck size. During an acute infection, mutations are constantly being introduced into the viral genome. Depending on timing of a mutation and its persistence, the relative frequency of each intrahost single nucleotide variant (iSNV) can vary. Like how barcodes are used in experimental systems, inference methods can use these iSNV frequencies to estimate the transmission bottleneck. Multiple methods exist to estimate bottleneck size. McCrone et al. used an early method that relies on the presence or absence of iSNVs between two members of a transmission pair (10). While conceptually simple, this method excludes donor iSNV not found in the recipient host at time of sampling and fails to account for stochastic nucleotide variant dynamics in early infection following transmission. Sobel Lenoard et al. subsequently developed the beta-binomial model that utilizes relative frequencies of shared iSNVs to estimate bottleneck size (11). The beta-binomial model was enhanced by Ghafari et al. to explicitly account for non-independent assortment of iSNVs during transmission (12). All these methods use frequencies of iSNVs (i.e., sites at which two alleles, or nucleotide variants, are present). One limitation, then, is that iSNV inclusion is strongly influenced by the nucleotide variant calling threshold, or the cutoff value determining the potential of a true genetic variation. With this in mind, the clonal mutation model developed by Shi et al. instead relies on consensus sequence differences between members of a transmission pair (13). This approach circumvents the limitations of the nucleotide variant calling threshold and better accounts for early infection stochastic effects.

Nearly all published data on the influenza A virus bottleneck originate from a single household study with 43 transmission pairs (14). While limited in the number of pairs, the estimate for this dataset is well validated with multiple studies finding that, regardless of inference model used, the transmission bottleneck is tight (1-3 virions) (11,13,15). However, given that only a single dataset has been used, these studies have not been able to assess the impact of host and viral factors on bottleneck size. This is a significant knowledge gap, as previous work has shown that influenza vaccination, influenza season, and viral subtype influence shedding and the secondary attack rates (16). Further, host factors such as sex at birth, age, and immune status are well documented modulators of viral shedding and transmission (17–19). Given their strong influence on transmission dynamics, we hypothesize that these same factors may also play a role in the transmission bottleneck size. Nonetheless, the impacts of viral and host factors on the transmission bottleneck for influenza, or many other viruses, are largely unknown.

Here we use data and specimens from two case-ascertained household transmission studies to determine how the influenza virus transmission bottleneck is modulated by viral and host factors. Specifically, we identify within-household transmission pairs and employ both the beta-binomial and clonal mutation models to estimate transmission bottleneck size. Each model utilizes distinct aspects of the viral sequence data, and, to our knowledge, this study compares these models on the largest dataset to date. Lastly, we examine how viral (season and subtype) and host factors (sex, age, and vaccination status) impact the transmission bottleneck size by examining subsets of our overall population stratified by these factors.

## METHODS

### Cohort description

We used data and specimens collected in two case-ascertained household transmission studies (FluTES and RVTN-S) based in the United States (US) that together spanned four influenza seasons. FluTES enrolled individuals from the 2017/2018, 2018/2019, and 2019/2020 seasons, and RVTN enrolled individuals in the 2021/2022 season. Both studies followed a similar design, and complete study protocols are published elsewhere (20,21). Briefly, enrollees were identified and recruited from ambulatory clinics, emergency departments, or other settings that performed influenza testing and received a positive test result for influenza A virus (IAV). Cases with acute illness of less than 5 days’ duration who lived with at least 1 other person who was not currently ill were eligible to participate. The enrollee and their household contacts were enrolled within 7 days of the enrollee’s illness onset and followed for up to 7 days (2017–2020) or 10 days (2021–2022). Nasal swabs were self-/parent-collected or staff-collected daily during follow-up and tested for influenza using RT-qPCR. Enrollment questionnaires were administered by study staff or self-administered. Questionnaires included information on participant age, self-reported sex and race, self-reported ethnicity, household characteristics, self-reported medical history, self-reported symptoms in the week prior to enrollment, and self-reported influenza vaccination for the current season. Influenza vaccination was self-reported at enrollment and was included if both date and location of vaccination were provided. Participants who reported vaccination less than 14 days prior to enrollment or who reported unknown vaccination were considered unvaccinated. Self-administered daily quantitative symptom questionnaires were completed during follow-up. These studies were approved by the Vanderbilt University Medical Center Institutional Review Board (FluTES IRB #171420, RVTN-S IRB #211495), reviewed by CDC, and conducted consistent with applicable federal law and CDC policy (45 CFR 46.102(l)(2)). All adults provided written consent; for children, parents provided written permission and children ≥ 7 years old provided assent.

### Sequencing of samples

We sequenced samples from households with two or more IAV positive individuals. We sequenced the first positive specimen with a cycle threshold (Ct) value of ≤30 from each individual to capture samples closest to likely time of transmission (22). IAV-positive samples with an RT-qPCR Ct ≤30 were sequenced in duplicate after the RNA extraction step. RNA was extracted using Invitrogen PureLink Pro 96 Viral RNA/DNA Purification Kit on an EpiMotion liquid handler or a MagMAX viral/pathogen nucleic acid purification kit (ThermoFisher) on a Kingfisher instrument. SuperScript IV one-step RT-PCR kits and universal IAV primers were used for RT-PCR (23). Sequencing libraries were prepared using Illumina DNA Prep Kits, and libraries were sequenced on a Novaseq (2 × 150 PE reads) by the Advanced Genomics Core at the University of Michigan or on a Nextseq (2 X 150 PE reads) in the Lauring laboratory. Reads from each sample were aligned to the corresponding influenza vaccine strain for each subtype and season: A/Michigan/45/2015 A(H1N1)pdm09, A/Hong Kong/4801/2014 A(H3N2), and A/Singapore/INFIMH-16-0019/2016 A(H3N2). For 2019/2020 A(H1N1)pdm09, we used A/New Jersey/13/2018, as the A/Brisbane/02/2018 sequence was not available. For the 2021/2022 influenza season we used A/Darwin/9/2021 A(H3N2). The vaccine strain for that season was A/Cambodia/e0826360/2020, but there was a mismatch between vaccine and circulating strain. For alignment, we used Bowtie2 (24) with the “very sensitive” setting, and duplicate reads were discarded using Picard tools (Picard Toolkit 2019). Reads from both replicates of a given specimen were combined and used to make a within-host consensus sequence using a script from (25). The replicates were then separately aligned to this consensus, and duplicates were removed.

### Nucleotide variant calling

For all genomes with an average genome-wide coverage of at least 1000× in both replicates (post de-duplication), we used iVar (26) to generate a consensus sequences and perform variant calling. We used an iSNV frequency threshold of ≥0.005 (0.5%). Intrahost single nucleotide variants had to be on reads with a mapQ of ≥20, bases with a phred score of ≥30, a per-site sequencing depth of ≥400, and an iVAR p-value of ≤1 × 10^−5^. iSNVs were retained only if they were called in both sequencing replicates. In the case of overlapping open reading frames (ORFs), an iSNV was classified as nonsynonymous if it was nonsynonymous in any ORF. Stop codons were classified as nonsynonymous. For all other analyses, we used the average iSNV frequency in the two replicates as the iSNV frequency. We excluded samples with greater than 50 iSNVs due to sequencing quality concerns. Further, as described in (11), iSNVs found only in recipient samples were excluded from analysis due to the low probability of the genetic difference emerging post transmission event.

### Clonal mutation determination

For the clonal mutation analysis, we counted the number of clonal mutations on a per-pair basis. Clonal mutations within a transmission pair are sites that (a) do not contain an iSNV and (b) have different nucleotides in each member of the transmission pair. To account for sequence degradation and artefacts typical to ends of IAV segments, we excluded any sites outside of the first start codon and last stop codon per segment.

### Delineation of transmission pairs

To generate phylogenetic trees, consensus sequences were aligned using MAFFT (v7) (27). Maximum likelihood phylogenetic trees were generated using IQTree (v2.4.0) with a GTR model (28). Trees were visualized and annotated using ggtree (29).

### Bottleneck size estimation

We utilized two methods to calculate transmission bottleneck size. The first method (11) uses iSNV data to determine pair-scale transmission bottleneck sizes. To obtain bottleneck estimates for the entire cohort or subgroup, we used an average weighted by the number of iSNVs per pair calculation (11). This method, as opposed to a simple average, accounts for differing number of iSNVs used in pair-scale transmission bottleneck estimates with pairs with greater number of iSNVs more heavily contributing to the cohort or subgroup transmission bottleneck estimate. The second method (13) uses clonal mutation distributions to determine transmission bottleneck sizes across groups of pairs. This method does not allow for pair-scale transmission bottlenecks to be calculated; instead, a distribution is fit to clonal mutation frequency data to estimate the bottleneck. For overall and subgroup analyses, separate distributions were generated and fit to obtain bottleneck estimates.

When iSNVs were utilized for bottleneck analysis, it was necessary to assess whether the consensus base at a particular locus was the same between both members of the transmission pair. In cases where both members of a pair contained iSNVs at a particular locus and, at that locus, they had the same consensus and alternative bases, iSNV frequencies were not altered. In most cases, only one member of a transmission pair contained an iSNV at a particular locus. If the consensus base was concordant (i.e., the same between both members of the transmission pair), the non-iSNV-containing member was assigned an iSNV frequency of 0. If the consensus base was discordant and the alternative base was the same as the consensus base in the opposite pair member, the iSNV frequency of 1 was assigned to the non-iSNV-containing member.

### Definition of cofactors

For secondary subgroup analysis, we used participant-reported metadata to define host cofactors. We defined vaccination as receipt of a season-specific influenza vaccine at least 14 days prior to enrollment. We defined sex based on subject self-report. We treated age as a dichotomous variable: individuals < 18 years as children and individuals ≥ 18 as adults. We subset the overall transmission pair population based on these defined cofactors within each role for pair members as described in **Results**.

### Data and code availability

Bottleneck estimation and all figures were generated using R (R version 4.4.2). Complete code to produce all paper figures can be found at https://github.com/lauringlab/IAV_bottleneck_Flutes_RVTN. Raw sequence reads are available on NCBI SRA, Bioproject PRJNA1085292 (FluTES) and PRJNA1303715 (RVTN-S).

## RESULTS

We obtained samples from individuals enrolled in FluTES and RVTN-S, two case-ascertained household transmission studies conducted in the United States. In full, these studies enrolled 2683 individuals from 806 households (20). In total, we sequenced 567 samples from 319 individuals and 102 households in duplicate. Of these, 399 were successfully sequenced (see Methods section for details).

We focused our analysis on samples and sequences surrounding transmission events. Households that contained viruses that differed by more than 5 mutations were excluded because this distance likely indicated multiple independent introductions rather than within-household transmission. A phylogenetic analysis confirmed the genetic relatedness of viruses within the remaining households (**Fig S1A-D**). Lastly, given the infectious period of IAV (30), we restricted our analysis to households in which symptom onset for all individuals within a household occurred within a continuous 14 day period. Following this filtering, we had a final analysis set of 100 participants from 46 households. Key descriptive features of included participants are shown in **Table 1**.

**Table 1.**
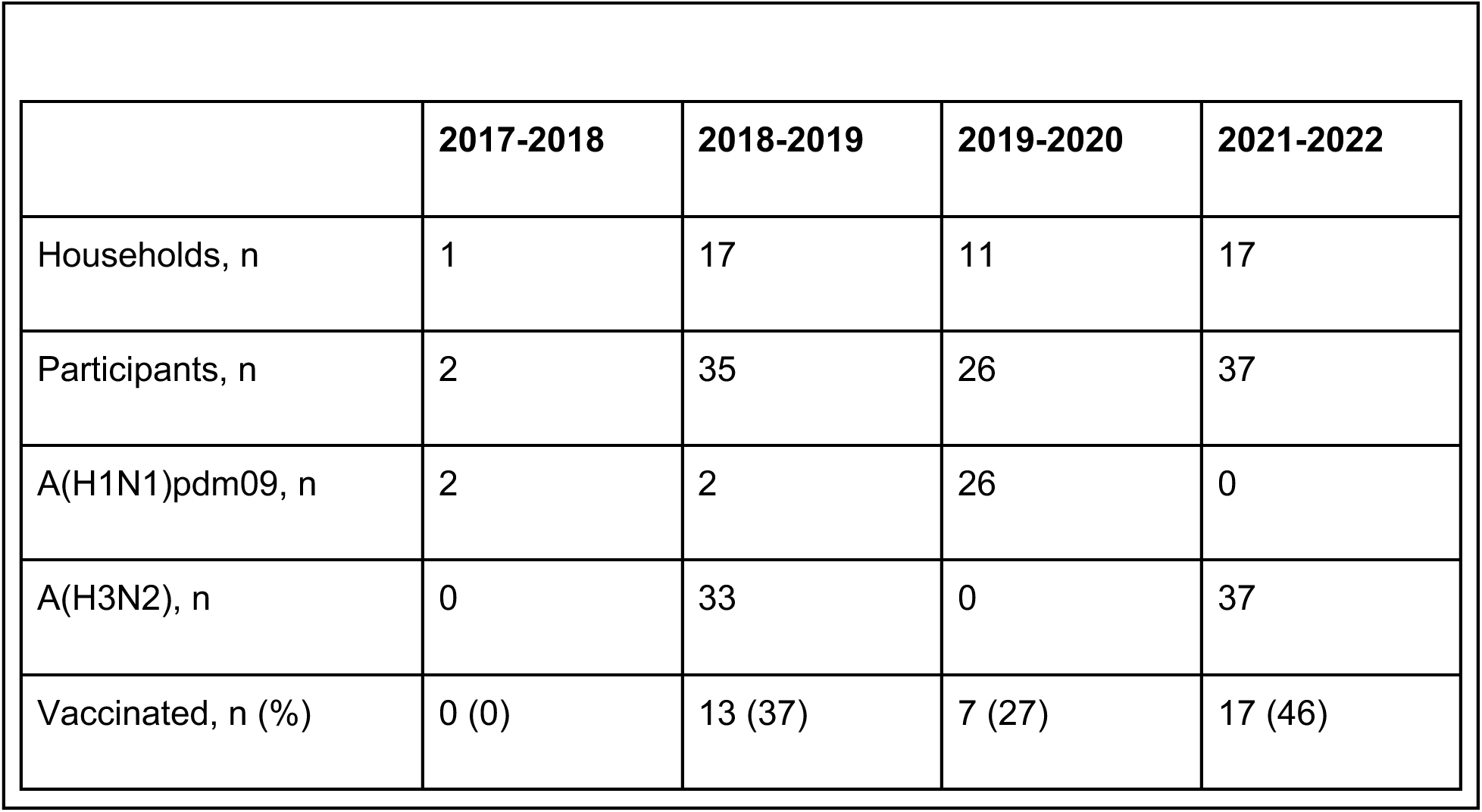
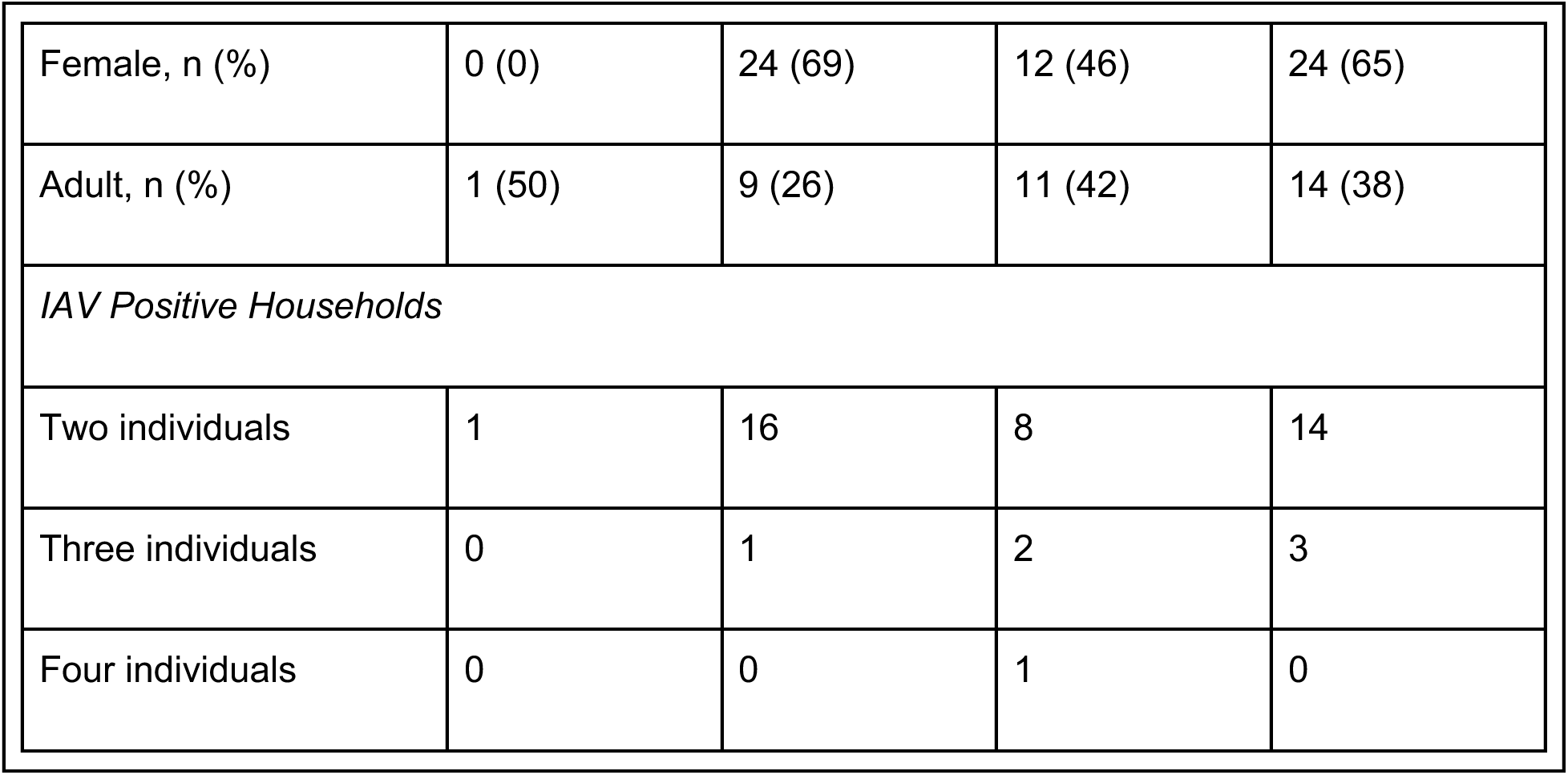
Influenza samples over four seasons within two household transmission studies.

Consistent with seasonal IAV trends in the United States, the A(H1N1)pdm09 subtype predominated during the 2019/2020 season while the A(H3N2) subtype predominated during the 2018/2019 and 2021/2022 seasons (31–33). The A(H3N2) subtype predominated in the 2017/2018 season (34); however, the single pair in this study had A(H1N1)pdm09. Regardless of season, most participants were unvaccinated or had an unknown vaccination status and a majority were under the age of 18 years. Aside from the 2019/2020 season, participants were primarily female. Most households had only two IAV-positive individuals with a handful of larger (3-4 person) households identified in each season.

Next, we examined genetic features of viruses included in our final analysis set. For all sequenced specimens, the frequencies (**Fig 1A**) and number (**Fig 1B**) of iSNV per sample were generally low, consistent with previous studies of IAV (35,36). We further found that iSNV frequency was strongly correlated between sequencing replicates (**Fig 1C**) indicative of high reproducibility in our variant calling and frequency estimates. There was no major difference in the iSNV frequency distributions of synonymous and nonsynonymous mutations, consistent with previous work (**Fig 1D**) (14,35).

**Figure 1.**
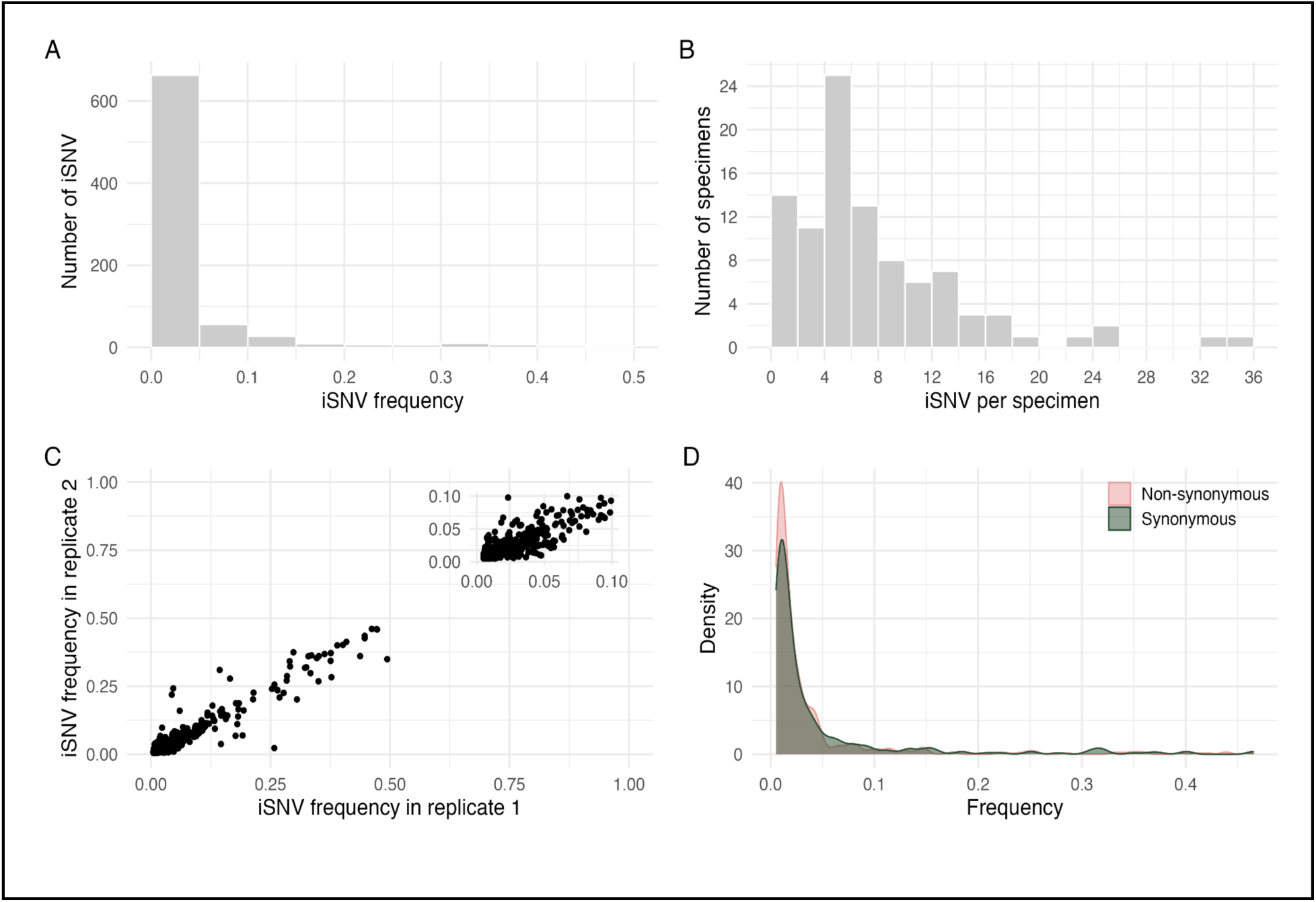
Genetic diversity of specimens. (A) histogram of iSNV frequency. (B) histogram of number of iSNVs per specimen. (C) scatter plot of iSNV frequency between sample replicates. (D) density plot of iSNV frequency of synonymous and non-synonymous mutations.

### Transmission pair characteristics

We defined transmission pairs from members within the same household and season based on symptom onset date. Most households had symptom onset of all individuals within one week (**Fig S2**). We assigned individuals with the earliest symptom onset date as donors for a household regardless of household size. For majority of cases, donor was study-identified index; in some cases, samples from study-identified index were not available/was not successfully sequenced and earliest symptom onset date was used to assign donor status. In cases of two individuals sharing an earliest-for-a-household symptom onset date, we treated both individuals as potential household donors and then as a donor-recipient pair with one another. In households with three or more individuals with unique symptom onset dates, we did not allow for individuals with intermediate onset dates to serve as donors. Our pairing method meant that some pairs were included twice to account for both directions of plausible transmission. For the clonal mutation analyses, in which directionality is not important, we excluded pairs that had identical individuals with directionality reversed (i.e., removed double counted pairs, see below). Additionally, in these cases, we assigned donor and recipient roles, however these role assignments were not relevant in the analysis. **Fig S3** shows a visual representation of this pairing schema.

Because the beta-binomial and clonal mutation models rely on different underlying sequence characteristics, the number of eligible pairs in our dataset differed. The beta-binomial model utilizes shared and non-shared iSNVs between members of a transmission pair and, thus, requires that samples contain at least one iSNV. This model is also sensitive to transmission direction. In contrast, the clonal mutation model utilizes the number of clonal differences between members of a transmission pair and, accordingly, does not impose any iSNV requirements for use of a sample. These differences in pairing requirement meant that the beta-binomial model analyzed more pairs, but fewer samples compared to the clonal mutation model.

**Table 2** describes characteristics of transmission pairs constructed for both methods of transmission bottleneck estimation.

**Table 2.** Transmission pair characteristics.

| Table 2. Transmission pair characteristics. |  |  |
| --- | --- | --- |
|  | Beta-binomial<br>method | Clonal mutation<br>method |
| Number of participants | 95 | 100 |
| Number of households | 44 | 46 |
| Number of possible transmission pairs | 59 | 56 |
| **Viral co-factors** |  |  |
| 2017-2018 | 1 | 1 |
| 2018-2019 | 22 | 19 |
| 2019-2020 | 16 | 15 |
| 2021-2022 | 20 | 21 |
| A(H1N1)pdm09 | 18 | 17 |
| A(H3N2) | 41 | 39 |
| **Host co-factors** |  |  |
| *donor characteristic / recipient characteristic* |  |  |
| (Unknown or influenza unvaccinated) / (unknown or influenza unvaccinated) | 31 | 28 |
| (Unknown or influenza unvaccinated) / influenza vaccinated | 5 | 5 |
| Influenza vaccinated / (unknown or influenza unvaccinated) | 8 | 9 |
| Influenza vaccinated / influenza vaccinated | 15 | 14 |
| Female / female | 21 | 17 |
| Female / male | 8 | 7 |
| Male / female | 21 | 24 |
| Male / male | 9 | 8 |
| Adult / adult | 5 | 5 |
| Adult / child | 12 | 9 |
| Child / adult | 17 | 19 |
| Child / child | 25 | 23 |

We evaluated transmission pairs that spanned our host and viral co-factors of interest. Aside from the 2017/2018 season where only two samples were included in our final analysis set, we had a similar number of pairs in each influenza season. We had a greater number of A(H3N2) transmission pairs as compared to A(H1N1)pdm09. This was attributable to a greater number of pairs within the A(H3N2)-predominated seasons (2018/2019 and 2021/2022) as compared to the A(H1N1)pdm09-predominated seasons (2017/2018 and 2019/2020). Our dataset contained more pairs with concordant vaccination status as compared to discordant vaccination status. We also found that female participants were represented more often in the recipient position as compared to the donor position. However, this was not relevant to transmission bottleneck estimation using the clonal mutation model as directionality does not contribute. Lastly, we found that transmission where at least one member of the pair is a child (under 18) was common in our population, reflective of our sample population containing more children than adults.

We next examined genetic features shared between members of a transmission pair. We found that the majority of iSNVs were not shared between members of a pair (**Fig 2A**). The number of clonal differences was also low for most transmission pairs (**Fig 2B**). These data are suggestive of a tight bottleneck.

**Figure 2.**
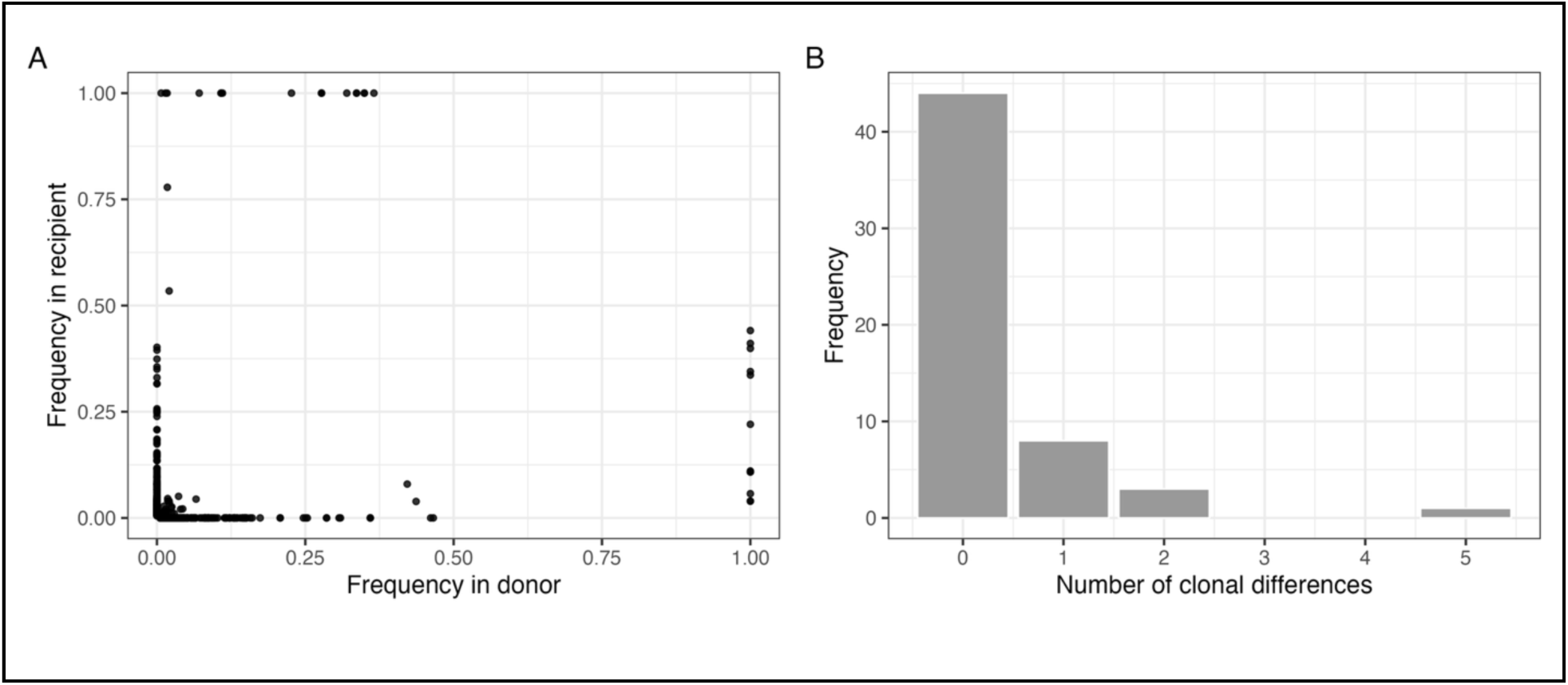
Shared iSNV and clonal distribution. (A) Shared genetic diversity between transmission pairs. Each point is an iSNV within a transmission pair. iSNVs are based on within host consensus sequence (≥ 50% frequency) and numerical corrections account for differences in between-pair consensus base at a particular locus. Details of numerical corrections applied can be found in **Methods.** (B) Distribution of number of clonal differences between transmission pair members for the overall study population.

### Transmission bottleneck size

To determine transmission bottleneck size, we employed two distinct approaches to utilize the two types of data available. Using the iSNV method (beta-binomial model), pair-scale transmission bottleneck estimates were heterogeneous in size (**Fig S4**). Generally, transmission bottleneck estimates for A(H1N1)pdm09 pairs were larger than for A(H3N2) pairs. When summed together using a weighted average method (see (11) for details), we found an overall transmission bottleneck size of 1. This is consistent with previous estimates using similarly structured datasets (37,38). The clonal mutation method does not permit pair-level estimates of transmission bottleneck size; nonetheless, it also estimated an overall transmission bottleneck size of 1 (exact estimate 0.91).

We next sought to determine if viral or host co-factors of the donor and/or recipient alter transmission bottleneck size. For viral co-factors, we evaluated season and viral subtype. For host co-factors, we evaluated age (child <18, adult ≥ 18), sex, and vaccination status. We found that stratification by host and viral factors did not strongly impact the estimated transmission bottleneck size regardless of the model used (**Fig 3**). In all cases when the beta-binomial model was utilized, we found that the transmission bottleneck of maximum likelihood was 1. Similarly, in all cases evaluated with the clonal mutation model, except for the 2017/2018 season, we found that the transmission bottleneck of maximum likelihood was 1 (exact estimate, 0.91). The 2017/2018 season had a slightly smaller estimate, and notably only had a single pair. Together, these data indicate that transmission bottleneck size is not strongly impacted by host or viral co-factors.

**Figure 3.**
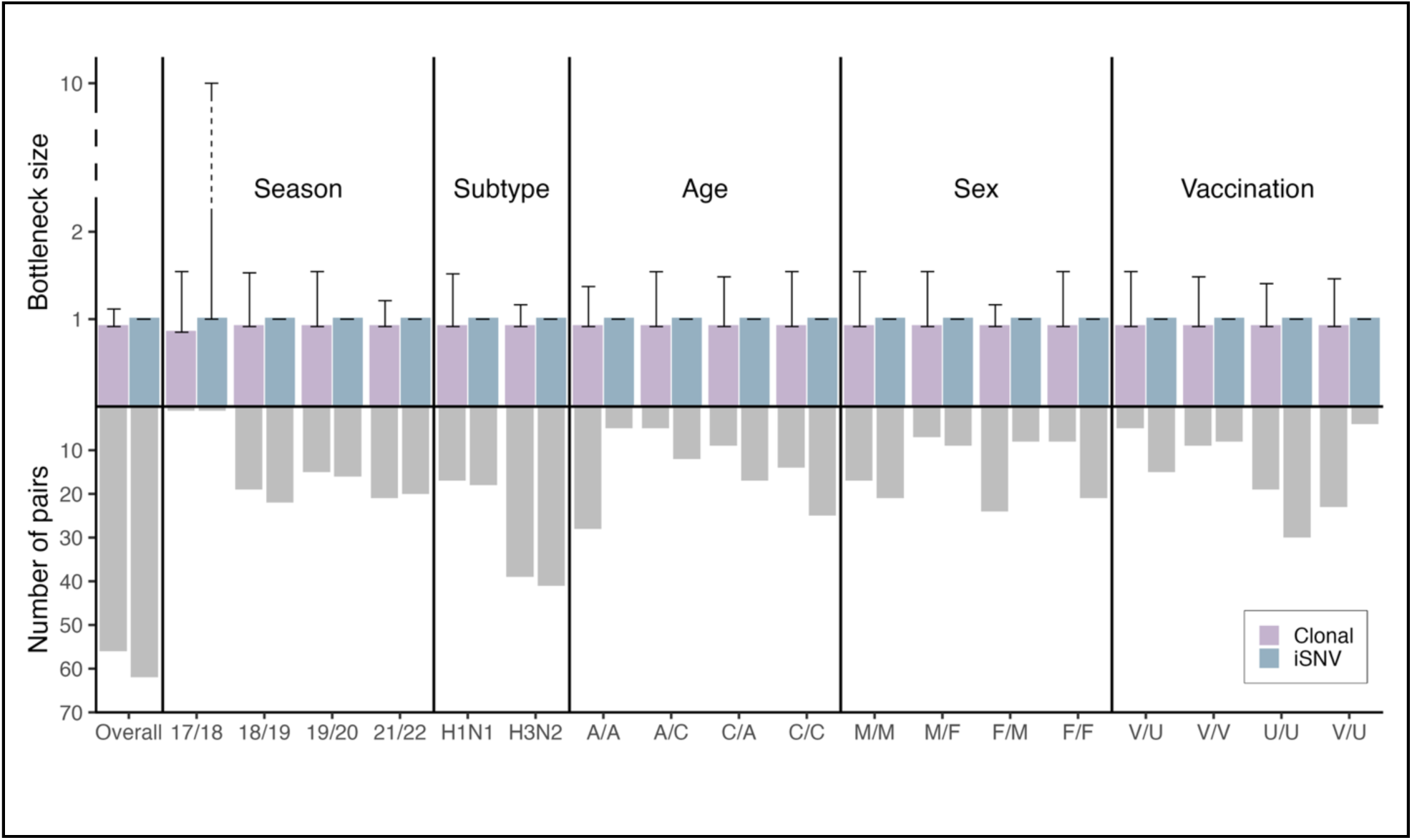
Bottleneck size with sample size overall and by metadata factors. Bottleneck estimates for overall population and by host/viral factors (year, subtype, age, sex, and vaccination status). Overall population and subgroup analysis bottleneck estimates were calculated with independent distributions (clonal method) or a weighted averaging method (iSNV method). Additional details on bottleneck estimation method can be found in **Methods**. Purple bars represent estimates found using clonal model and blue bars represent estimates found using beta-binomial model (in all cases, 1). Host factors are listed in format of X/Y with X representing donor factor and Y representing recipient factor. “A” stands for adult. “C” stands for child. “M” stands for male. “F” stands for female. “V” stands for vaccinated. “U” stands for unvaccinated or unknown vaccination status. 95% confidence intervals are shown by black error bars.

## DISCUSSION

In this study, we utilized two case-ascertained household transmission studies spanning 4 influenza seasons to estimate the transmission bottleneck across more than 50 possible household transmission pairs. We used two different estimation methods, each leveraging distinct features of sequencing data. We found that, regardless of method used, transmission bottleneck size was approximately 1 nucleotide variant, indicating a clonal viral population establishes infection. While factors such as influenza season, subtype, and influenza vaccination status, sex, and age (child vs. adult) are known to influence influenza viral shedding and transmission, they did not alter the estimated transmission bottleneck size in our population.

Influenza virus evolution has been studied extensively at both the host and population scales. Transmission is critical because it links these two scales – a nucleotide variant must be transmitted from one host to another to spread across a population. Bottleneck size is important as it determines how much diversity is lost during a transmission event. This study and others have found that the transmission bottleneck for influenza virus is very tight. Further, while some data are consistent with a contribution of positive selection to transmission (39), a majority of evidence points to stochastic effects determining the virions that survive the bottleneck (40); the frequency of a given nucleotide variant in the population of incoming infectious particles at the moment of exposure determines its likelihood of being successfully transmitted and, when the bottleneck is tight, typically the majority variant survives. Given that influenza viral shedding typically starts 2 days post infection (41), nucleotide variant frequency is largely set by the frequencies in the founding viral population. As newly arising nucleotide variants rarely accumulate in that short a time to a frequency where they can plausibly be transmitted, within-host processes are likely not a significant contributor to population level processes for IAV. In fact, for SARS-CoV-2 infection, studies show that evolution within a single host can outpace evolution along a transmission chain (8,42).

We also find that factors known to impact viral shedding and transmission rates do not impact transmission bottleneck size. For instance, Rolfes et al. performed an analysis on a subset of the same case-ascertained household cohorts we utilized and found a significantly higher secondary attack rate in the 2021/2022 IAV season compared to prior seasons (43). When comparing the same seasons, we did not find a corresponding difference in bottleneck size (**Fig 3**). This is not limited to influenza virus. Bendall et al. similarly found that the SARS-CoV-2 transmission bottleneck is not related to infection severity or transmissibility (22). Together, these findings suggest that a tight transmission bottleneck size may be intrinsic to the respiratory virus transmission route. Further, while barcoded systems have been used in animal models to show that transmission routes can alter the bottleneck size, tight bottlenecks have been consistently found in non-respiratory viruses (38,44–46). This suggests a tight bottleneck may be an essential feature of virus transmission. One hypothesis as to why this is the case is that there is simply a very small probability of successful infection establishment by multiple nucleotide variants. Respiratory virus transmission is a multistep process: a virion must evade the intrinsic and innate immune factors of the respiratory tract within its initial host, transit through the host respiratory tract, survive outside the body (as a droplet in the air or on a living (e.g., fingers) or inanimate (i.e., fomite) surface), enter a recipient’s respiratory tract, find recipient cells supportive of replication, and, finally, evade within-cell defenses of the recipient to successfully replicate. The chance that all these events will occur successfully for a single virion is very small. Where virions get lost along this pathway can be deciphered using barcoded systems, an approach that has been successfully used within animal models (39,47).

Our study has several strengths. To our knowledge, this is the largest number of high-quality transmission pairs used to estimate transmission bottleneck size from a single dataset. Further, primary sequence data underwent rigorous screening procedures. Our study used first positive samples obtained over the course of daily monitoring. This means that there is a high likelihood that we accurately capture viral heterogeneity very close to time of transmission. Given that transmission bottleneck estimation involves using inference models to reconstruct the genetic heterogeneity at the precise moment of transmission, having samples with granular information on timing of infection pairs is ideal. We also employed two methods that have both been previously validated on similarly structured datasets and, as described in (11) and (13), are orthogonal approaches that utilize distinct aspects of sequence data to estimate transmission bottleneck size.

Our study also has some limitations. As is the case with all human datasets, our study may not be completely generalizable. Our study population represents a relatively healthy population and does not cover settings where persistent viremia and transmission potential are more prevalent (e.g., those in residential/long-term care). Further, factors not controlled for such as socioeconomic status, race, air quality, or climate may mean that our findings are not valid outside of the study population.

Due to our approach to identifying transmission pairs, we include some pairs twice to account for uncertain transmission directionality. While this slightly dilutes our sample set, we believe that it allows for increased certainty, as we capture true directionality of transmission. We have also potentially included transmission pairs that are not valid. Given typical social patterns of households, there is potential for two household members to have joint exposure to an external source as opposed to our assumption that one household member always transmits to another household member. We mitigated this by requiring minimal genetic distance between pairs prior to analysis. We dichotomize age into two categories, adult and child, as opposed to further dividing age based on known lines of differing immune differences (e.g., delineating older vs. younger adults) (48). While this limits the scope of questions addressed, further divisions of age categories would decrease our sample size for each subgroup. Lastly, some of our population subsets and co-factor analysis groups are small, limiting the statistical power of our study. However, we are confident we could detect consequential differences in transmission bottleneck size if they were to exist.

We find that the transmission bottleneck size of influenza A virus is small regardless of viral and host cofactors known to modulate intra- and inter-host infection processes. Given a strong role for antigenic drift in IAV evolution, our finding that the transmission bottleneck is tight raises interesting questions regarding how within-host and population-scale IAV evolution are linked mechanistically, culminating in well-established paradigms of influenza ecology and epidemiology. Ultimately, our findings suggest that there is a deeper genetic or mechanistic basis for transmission bottleneck size – the investigation into which will be the subject of future work.

## ACKNOWLEDGEMENTS

We thank all individuals who participated in this study. KK was supported by the Molecular Mechanisms in Microbial Pathogenesis Training Program, NIH T32 AI007528. The FluTES study was supported by CDC U01IP001083 (to HKT, CGG) and the RVTN-S study was supported by CDC 75D30121C11656 (to HKT, CGG). Sequencing and analysis in the Lauring lab was supported by NIH R01 AI148371 (to ASL, ETM) and Penn-CEIRR, NIH 75N93021C00015 (to ASL, ETM).

## AUTHOR CONTRIBUTIONS

Conceiving and Designing the Study: Lauring, Grijalva, Talbot Acquiring the Data: All authors

Analysis and Interpretation of the Data: Bendall, Krupinsky, Lauring Drafting the Manuscript: Krupinsky, Lauring

Editing the Manuscript: All authors

Supervising the Research: Lauring, Grijalva, Talbot Obtained Funding: Lauring, Martin, Grijalva, Talbot

The findings and conclusions in this report are those of the authors and do not necessarily represent the official position of the Centers for Disease Control and Prevention.

## SUPPLEMENTAL FIGURES AND TABLES

**Supplemental Figure 1.**
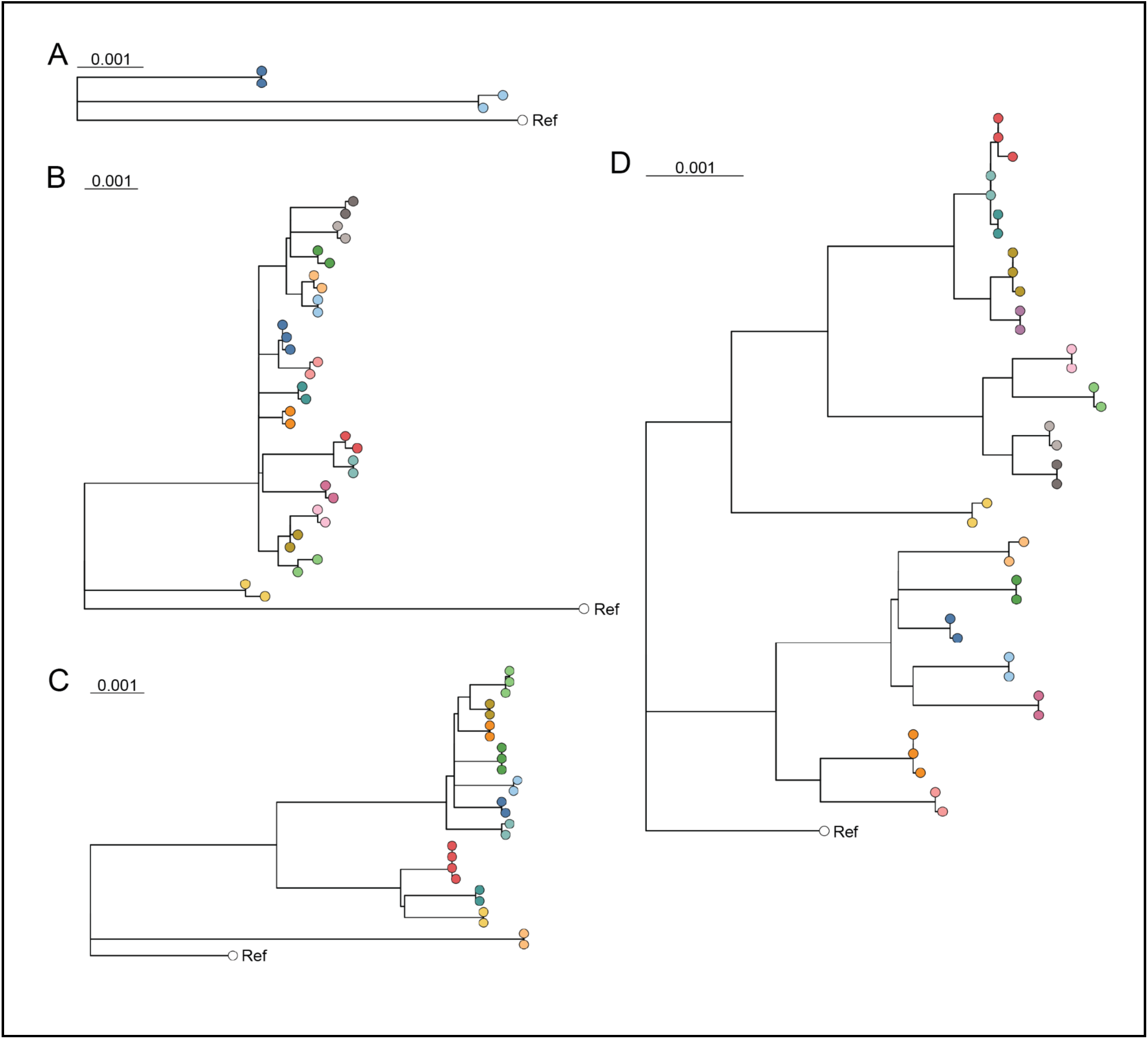
S**a**mple **phylogenetic trees.** (A-D) Phylogenetic trees of all samples. Tips are colored based on household membership with colors in separate trees representing non-related households. Individual panels represent unique reference strains as follows: (A) A/Michigan/2017 A(H1N1)pdm09, (B) A/Singapore/2018 A(H3N2), (C) A/Brisbane/2019 A(H1N1)pdm09, (D) A/Darwin/2021 A(H3N2).

**Supplemental Figure 2.**
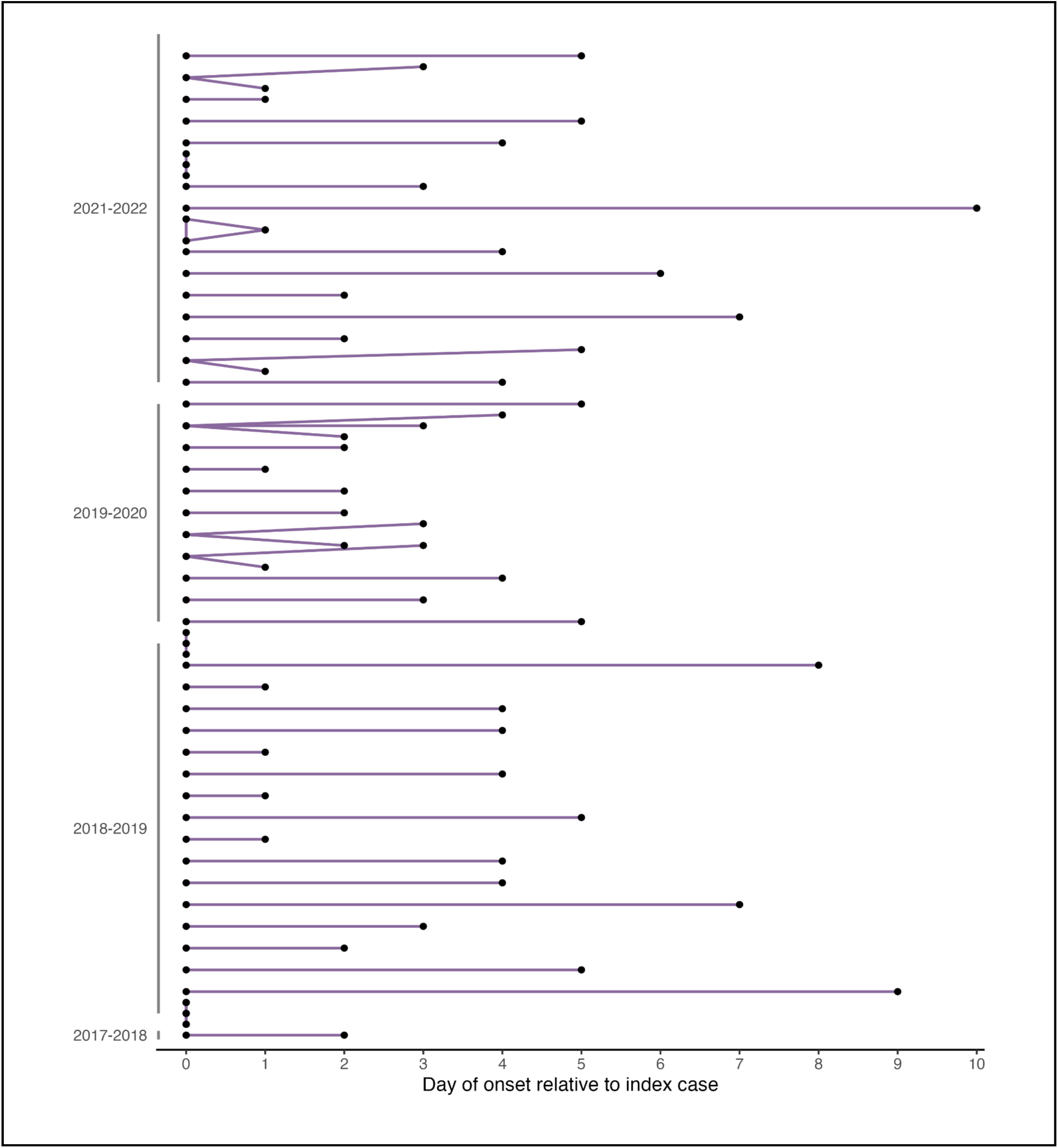
T**i**ming **of each pair relative to the index case.** Dots represent all individuals included in our final analysis set. Lines connect members of a transmission pair. Individuals with the earliest symptom onset date are defined as the index case. In cases where two individuals in a single household have an earliest-for-household onset date, both cases are assigned as index cases.

**Supplemental Figure 3.**
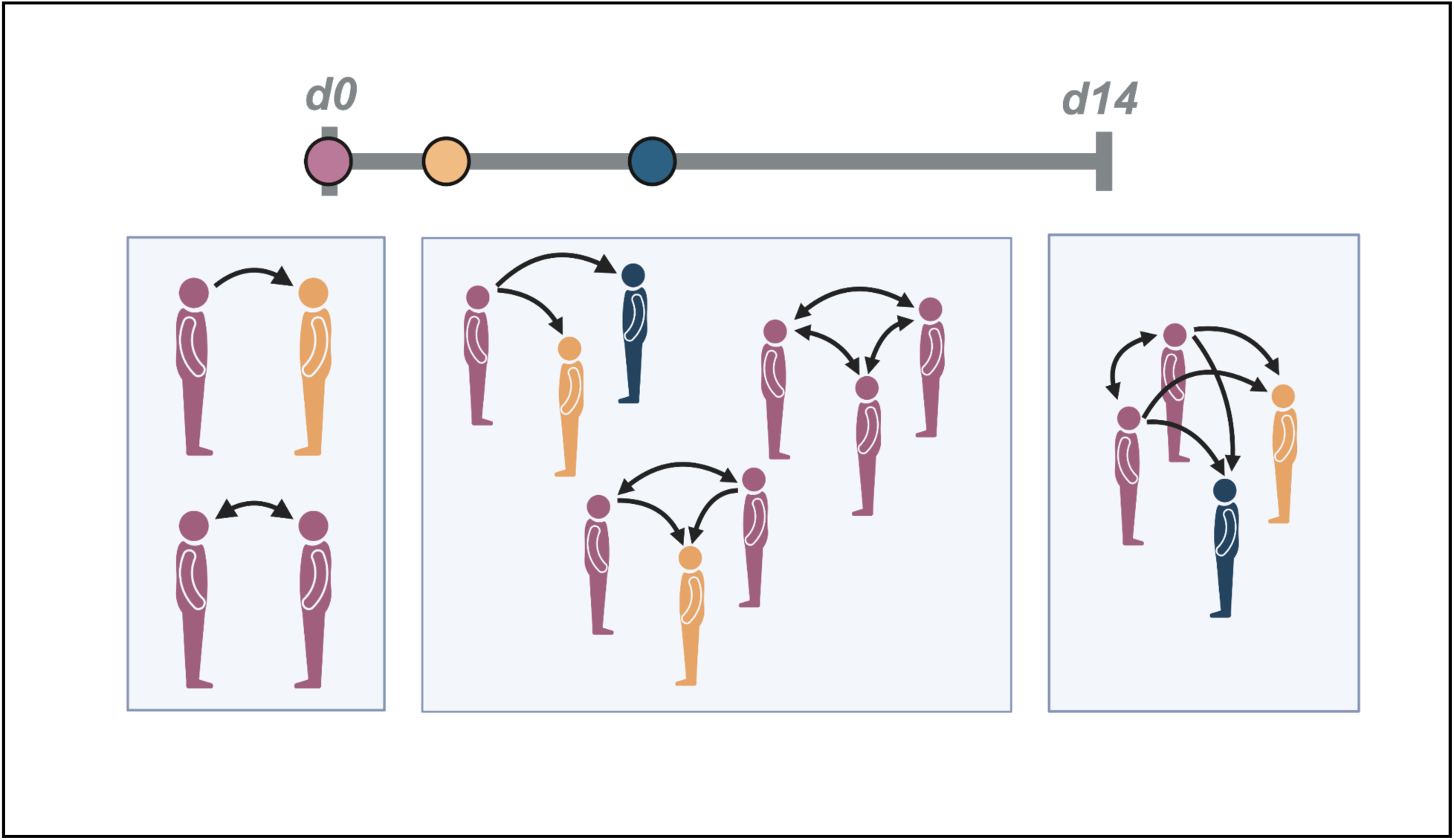
T**r**ansmission **pairing schema.** Individual samples are assigned into pairs based on the date of symptom onset. Each arrow indicates a transmission pairing with the arrowhead pointing towards a recipient.

**Supplemental Figure 4.**
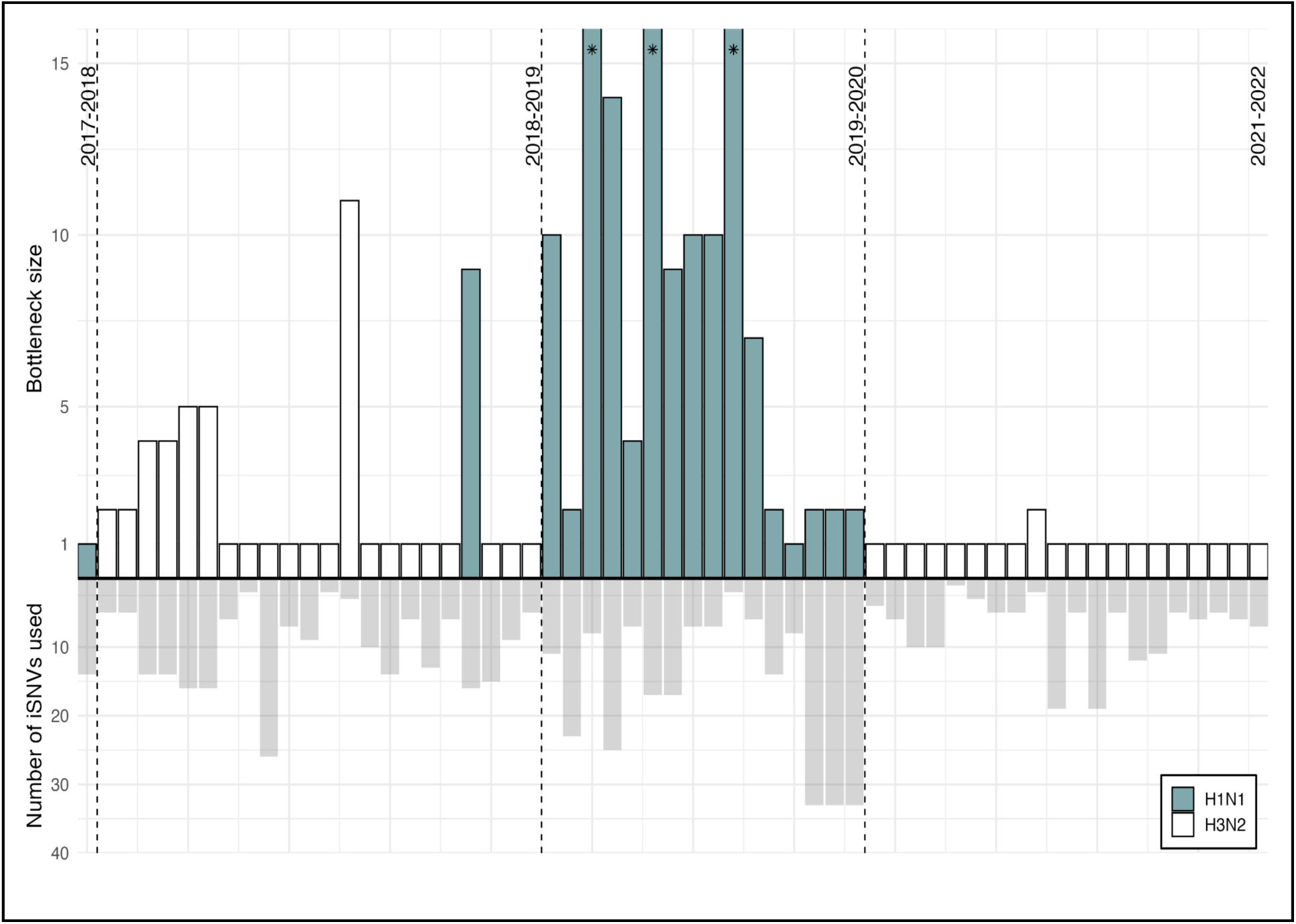
I**n**dividual **pair bottlenecks.** Bottleneck estimates for individual transmission pairs using beta-binomial model (top) and number of iSNVs used in each estimate (bottom). Filled in (teal) bars represent pairs with A(H1N1)pdm09; open (white) bars represent pairs with A(H3N2). iSNVs were used if they were found in the donor pair-member or both pair-members (i.e., iSNVs were not used if found exclusively in recipient pair-member). Stars on bars represent maximum likelihood estimates greater than 15.

